# Impact of relaxing Covid-19 social distancing measures on rural North Wales: a simulation analysis

**DOI:** 10.1101/2020.05.15.20102764

**Authors:** Rhodri P Hughes, Dyfrig A Hughes

## Abstract

**Background:** Social distancing policies aimed to limit Covid–19 are gradually being relaxed as nationally reported peaks in incident cases are passed. Population density is an important driver of national incidence rates; however peak incidences in rural regions may lag national figures by several weeks. We aimed to forecast the impact of relaxed social distancing rules on rural North Wales.

**Methods:** Daily data on the deaths of people with a positive test for Covid–19 were obtained from Public Health Wales and the UK Government. Sigmoidal growth functions were fitted by non-linear least squares and model averaging used to extrapolate mortality over time. The dates of peak mortality incidences for North Wales, Wales and the UK; and the percentage predicted maximum mortality (as of 7^th^ May 2020) were estimated.

**Results:** The peak daily death rates in Wales and the UK were estimated to have occurred on the 14/04/2020 and 15/04/2020, respectively. For North Wales, this occurred on the 07/05/2020, corresponding to the date of analysis. The number of deaths reported in North Wales represents 31% of the predicted total cumulative number, compared with 71% and 60% for Wales and the UK, respectively.

**Conclusion:** Policies governing the movement of people in the gradual release from lockdown are likely to impact significantly on areas –principally rural in nature– where cases of Covid–19, deaths and immunity are likely to be much lower than in populated areas. This is particularly difficult to manage across jurisdictions, such as between England and Wales, and in popular holiday destinations.

## Introduction

The Severe Acute Respiratory Syndrome Coronavirus-2 (SARS-CoV-2) has resulted in 4.17m cases of Covid-19 worldwide (as of 13^th^ May 2020) [1]. Declared a pandemic by the World Health Organisation in February the 11^th^ 2020, measures to contain the spread of SARS-CoV-2 has seen most countries impose social distancing measures including restrictions on travel, work and closure of non-essential services. On the 23^rd^ of March a lockdown was introduced in the United Kingdom (UK) to limit further spread of the virus.

These measures are likely to have had a significant impact on the number of daily confirmed cases of Covid-19 and deaths associated with Covid-19 infection. In the UK, the Prime Minister, Boris Johnson, announced some easing of the lockdown measures for England on the 10^th^ May 2020. However the guidance lacked some clarity on the implications to other countries within the UK. With devolved powers to enforce measures to control movement of people in response to Covid-19, the governments of Wales and Scotland have retained more strict social distancing measures.

Differences in policies between countries within the UK reflect geographical differences in disease incidence, prevalence and the reproduction number, R, which is estimated to be between 0.5 and 0.9 across the UK, nearer to 1 in Scotland, and 0.8 in Wales (10^th^ May 2020) [2]. Within Wales, the incidence of Covid-19 has varied substantially, with 446 cases per 100,000 in the South East (more populated, urban areas) to 247 cases per 100,000 in the North (sparsely populated and more rural) [3]. Policies driven by changes in transmission in populated areas (which have mainly peaked) may not be applicable to rural areas (where cases may yet to peak).

North Wales is primarily a rural region without any large cities. The north-east is more industrial and the majority of the population reside along the coastal areas. The north-west is sparsely populated (<50 people /km^2^), reliant on the tourism and agricultural economies. As a popular holiday destination, over 3.9m people visited the Snowdonia National Park alone in 2015 [4]; and there are more than 5000 second homes in north west Wales, where 1 in 3 properties are sold to residents from outside the region. North Wales is served by a unitary health authority (Betsi Cadwaladr University Health Board, BCUHB), providing primary, secondary, community and social care to 696,300 inhabitants. Increases in the population numbers risk placing pressure on the 3 district general hospitals that have 31 intensive care beds. In response to Covid-19, however, an additional 930 bed spaces have been made available via regional temporary hospitals.

During the weekend prior to the lockdown (21–22 March 2020) record numbers of tourists were reported to visit Snowdonia. The Snowdonia National Park Authority described an “unprecedented scene” which saw hundreds of people walking up Wales’ highest mountain in what the authority said was “the busiest visitor day in living memory” [5]. During this period there was also a surge in the number of people relocating – mainly from metropolitan areas of England – to their second homes in North Wales. In the days immediately following the easing of the lockdown in England (13^th^ May 2020), there were reports of holiday parks being “flooded” with booking requests, despite more strict laws applying in Wales [6].

The aim of the present analysis was to assess whether the trajectory of Covid-19 related mortality rates reported in BCUHB mirror those for Wales, and UK as a whole. Differences in the rate of increase of deaths, timing of peak rates and decline may indicate that different policies need to be enforced to limit transmission, pressures on the health service, and mortality.

## Methods

### Data

Daily data on the deaths of people with a positive test for Covid-19 were obtained from Public Health Wales [3] and the UK Government [7]. Both datasets include patients who may have died from other causes, and exclude the deaths of people who were not tested, or who might have died from (or with) Covid-19 but did not tested positive.

Data for the UK and Wales were obtained from the 08/03/2020 and 18/03/2020, respectively, to the 07/05/2020. Data for BCUHB were obtained between the 20/03/2020 and the 07/05/2020; however, daily data for BHUHB were missing between 21/03/2020 to the 23/04/2020 because of a data reporting error and the Health Board reported all of the deaths between these dates on the 24/04/2020. Prior to 21/03/2020, there were fewer than 5 cases of deaths, this being the threshold for disclosing information to avoid de-anonymization.

### Analysis

Missing daily data for BCUHB were imputed using the predictions from an exponential function fitted to observed data points. This expression was assumed to be applicable for historic data during the exponential growth phase of transmission. Nowcasts and forecasts of cumulative mortality were made using a range of sigmoidal growth functions: logistic, S-Shape, Richards, Weibull and Gompertz functions, which are defined below:

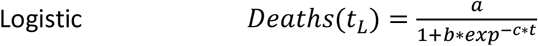

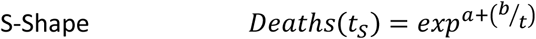

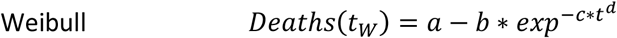

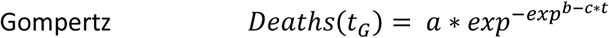

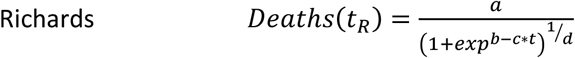

Each were fitted to the data by least squares using the non-linear regression function (CurveFit) in Stata version 13 (StataCorp, College Station, TX) [8] to estimate parameters a, b, c, d. Modelling uncertainty was considered using unweighted model averaging. The date of peak rate of deaths was estimated to correspond to the steepest incline in the rate of cumulative deaths. The maximum cumulative number of deaths was estimated for each region (BCUHB, Wales, UK), and the number of deaths to 07/05/2020 was expressed as a percentage of the predicted maximum number of deaths for each region of interest.

## Results

Figure 1 presents all the recorded daily deaths for the regions of interest. Convergence in the non-linear curve fitting was achieved for all functions other than Richards, which is equivalent to the Gompertz model when d approaches 0. The model parameter estimates, presented in Table 1, and resulting growth curves depicted in Figure 2 indicated the peak daily death rate in Wales occurred on the 14/04/2020 (range 11/04/2020 to 15/04/2020). Peak daily deaths for the UK occurred on 15/04/2020 (range 12/04/2020 to 20/04/2020) – both indicating that the first peaks for daily deaths have passed. For BCUHB, the peak for daily deaths occurred on the 07/05/2020 (range 02/05/2020 to 26/05/2020). This corresponds to the latest date for which data were available at the time of analysis, meaning that the date of peak daily deaths for BCUHB is highly uncertain, and may not yet have occurred.

**Figure 1.**
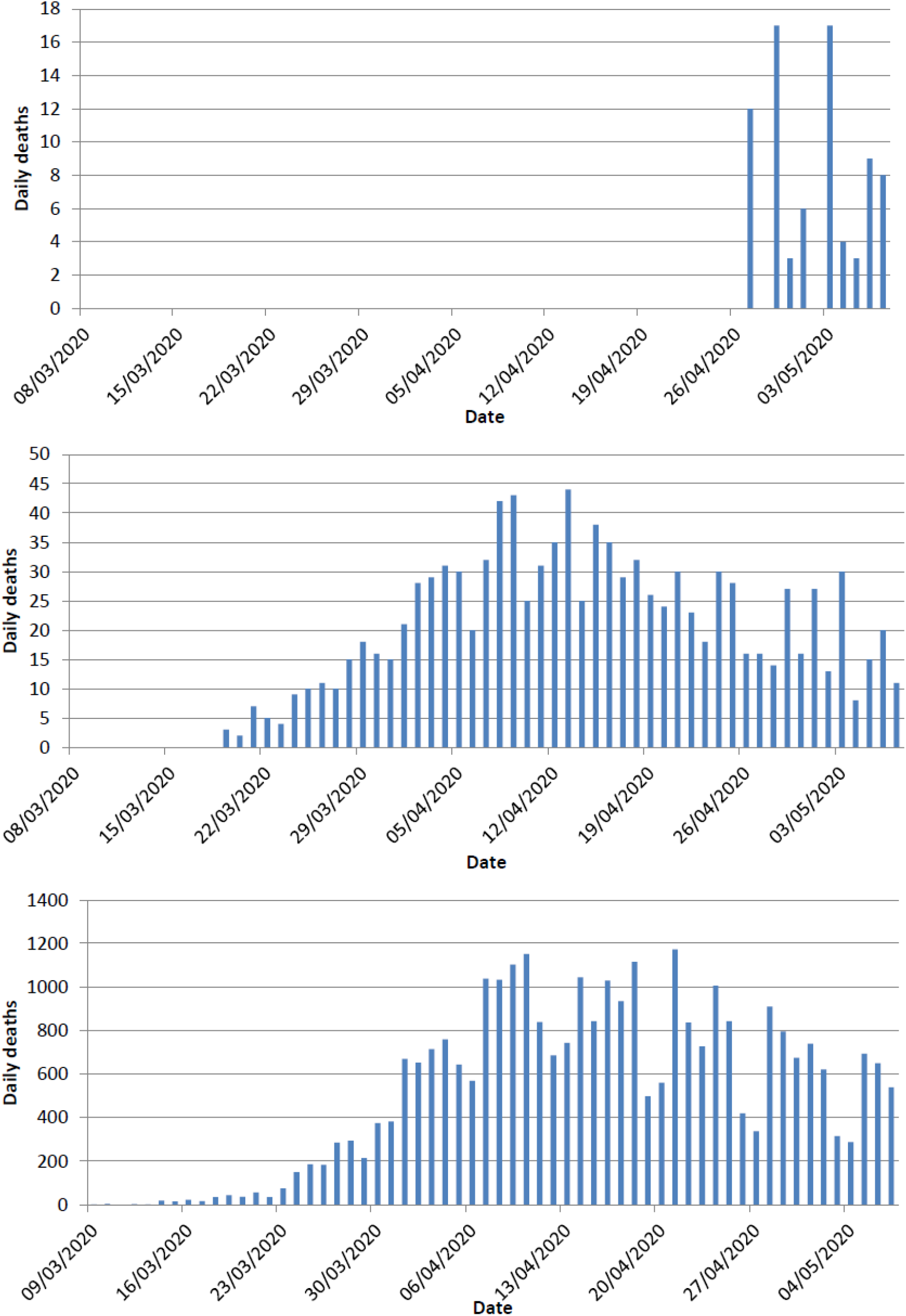
Frequency of deaths in BCUHB (top), Wales (middle) and the UK (bottom)

**Table 1.** Parameter estimates for each model. Data in parentheses are the standard errors.

| Model | a | b | c | d | r<sup>2</sup> |
| --- | --- | --- | --- | --- | --- |
| BCUHB |  |  |  |  |  |
| Logistic | 268.37 (16.38) | 214.94 (8.22) | 0.10 (23.46) |  | 1.000 |
| S-Curve | 7.44 (74.69) | -142.46 (-25.55) |  |  | 0.999 |
| Weibull | 266.22 (6.96) | 260.72 (6.66) | 0.00 (1.05) | 4.00 (14.12) | 0.999 |
| Gompertz | 779.23 (3.63) | 2.12 (38.98) | 0.03 (7.58) |  | 0.999 |
| Richards | 261.51 (4.44) | 5.67 (2.20) | 0.10 (2.48) | 1.07 (1.78) | 1.000 |
| Wales |  |  |  |  |  |
| Logistic | 1091.76 (87.69) | 172.05 (9.05) | 0.14 (39.03) |  | 0.999 |
| S-Curve | 8.28 (281.45) | -75.39 (-53.85) |  |  | 0.998 |
| Weibull | 1112.74 (93.47) | 1141.45 (71.94) | 0.00 (3.97) | 3.18 (45.04) | 0.999 |
| Gompertz | 1252.46 (166.53) | 2.56 (122.42) | 0.07 (95.01) |  | 1.000 |
| Richards | 1252.33 (166.62) | -5.20 (n/a) | 0.07 (95.08) | 0.00 (47.81) | 1.000 |
| UK |  |  |  |  |  |
| Logistic | 30984.59 (77.54) | 229.88 (8.55) | 0.14 (38.32) |  | 0.998 |
| S-Curve | 11.78 (425.55) | -86.02 (-63.81) |  |  | 0.998 |
| Weibull | 164.84 (3.77) | -53188 (-48.48) | 1130.74 (8.54) | -1.85 (-51.00) | 1.000 |
| Gompertz | 36523 (132.65) | 2.65 (112.48) | 0.07 (84.31) |  | 1.000 |
| Richards | 36521 (132.66) | -5.85 (n/a) | 0.07 (84.33) | 0.00 (42.46) | 1.000 |

**Figure 2.**
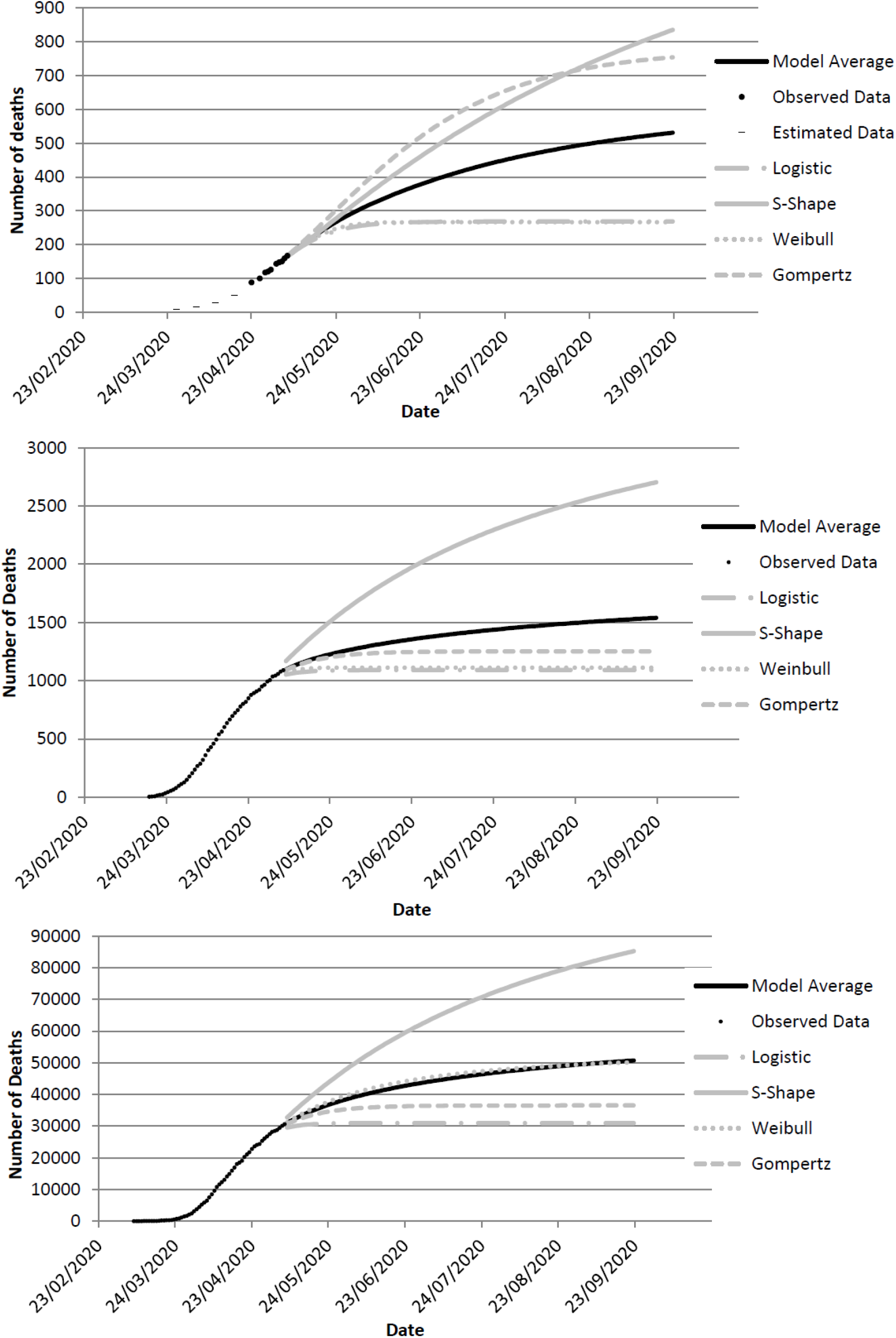
Observed and forecasted cumulative mortality in Covid-19 positive patients in BCUHB (top), Wales (middle) and the UK (bottom)

The final estimation of predicted deaths for the UK, Wales and BCUHB are, respectively, 51,000 (range 31,000 to 85,000), 1,540 (1,090 to 2,700), and 530 (270 to 835). As of 07/05/2020, the number of deaths reported for BCUHB (167) represents 31% of the predicted total cumulative number (range 6% to 63%), suggesting that the region is not yet halfway in terms of absolute numbers of deaths in Covid-19 positive patients. By contrast, deaths across Wales was predicted to be 71% (range 40% to 100%) of the total, and the UK 60% (range 36% to 99%).

## Discussion

The results suggest that the rate of Covid-19 positive deaths in Wales and the UK have peaked, although there is predicted significant mortality in the coming weeks and months, consistent with multiple other forecast models of Covid-19 [9]. The situation is different in North Wales, where there remains significant uncertainty concerning the timing of peak mortality. Concerns that the incidence of new cases may be rising at a higher rate than the remainder of Wales, and the ≥2 week lag in mortality implies that reducing current controls on population movement may be detrimental to the region’s population health.

The fragility of rural North Wales in dealing with Covid-19 in the context of substantial increases in holidaymakers and second home residents is significant. The May 10^th^ announcement of the relaxation in the lockdown for England, included freedom for exercise and outdoor activity, “irrespective of distance”. Whether this includes travel to Wales and other parts of the UK is unclear – the Welsh Government has ruled that stopping people breaking Welsh coronavirus lockdown laws is not a “real option”.

Our analysis has strengths in consideration of multiple sigmoidal growth functions, contrasting with many others, including the influential Institute for Health Metrics and Evaluation modelling which relies on a single model, namely the ERF error function. Their approach has been criticised as predictions are extremely labile since new data are included on a daily basis [10]. Neither our model nor the IHME model is a disease transmission model, and this represents a limitation. Although in predicting mortality (as opposed to cases), SEIR compartmental models (representing susceptible, exposed, infectious, recovered) may be less reliable. Model averaging benefits from possible reduction of predictive error. However the confidence bounds for averaged models are not readily calculable, hence our presentation of the range of outputs from each individual model as a conservative estimate. A further limitation relates to the data, as not all Covid-19 deaths are reported in NHS and Government figures. Estimations of excess mortality in relation to historic data for the months of March to May should provide a more robust estimate, and which are inclusive also of wider impacts of hospital pressures and cancellation of elective procedures.

In conclusion, policies governing the movement of people in the gradual release from lockdown are likely to impact significantly on areas – principally rural in nature – where cases of Covid-19, deaths and immunity are likely to be much lower than in populated areas. This is particularly difficult to manage across jurisdictions, such as between England and Wales, and for popular holiday destinations.

## Data Availability

Data are available from publicly accessible sources

https://public.tableau.com/profile/public.health.wales.health.protection#!/vizhome/RapidCOVID-19virology-Public/Headlinesummary

https://coronavirus.data.gov.uk/

## Contribution

Rh.P.H. made substantial contributions to conception and design, acquisition of data, analysis and interpretation of data; drafted the article; and give final approval of the version to be published. D.A.H. made substantial contributions to conception and design, acquisition of data, analysis and interpretation of data; revise the article critically for important intellectual content; and give final approval of the version to be published.

